# Locus coeruleus pathology in progressive supranuclear palsy, and its relation to disease severity

**DOI:** 10.1101/2020.01.13.20016360

**Authors:** Sanne Simone Kaalund, Luca Passamonti, Kieren SJ Allinson, Alexander G Murley, Trevor W Robbins, Maria Grazia Spillantini, James B Rowe

## Abstract

The locus coeruleus is the major source of noradrenaline to the brain and contributes to a wide range of physiological and cognitive functions including arousal, attention, autonomic control, and adaptive behaviour. Neurodegeneration and pathological aggregation of tau protein in the locus coeruleus are early features of progressive supranuclear palsy (PSP). This pathology is proposed to contribute to the clinical expression of disease, including the PSP Richardson’s syndrome. We test the hypothesis that tau pathology and neuronal loss are associated with clinical heterogeneity and severity in PSP.

We used immunohistochemistry in *post mortem* tissues from 31 patients with a clinical diagnosis of PSP (22 with Richardson’s syndrome) and 6 control cases. We quantified the presence of hyperphosphorylated tau, the number of pigmented cells indicative of noradrenergic neurons, and the percentage of pigmented neurons with tau-positive inclusions. *Ante mortem* assessment of clinical severity using the PSP rating scale was available within 1.8 (±0.9) years for 23 patients.

We found an average 49% reduction of pigmented neurons in PSP patients relative to controls. The loss of pigmented neurons correlated with disease severity, even after adjusting for disease duration and the interval between clinical assessment and death. The degree of neuronal loss was associated with tau-positive inclusions, with an average of 44% of pigmented neurons displaying tau-inclusions.

Degeneration and tau pathology in the locus coeruleus are related to clinical heterogeneity of PSP. The noradrenergic deficit in the locus coeruleus is a candidate target for pharmacological treatment. Recent developments in ultra-high field magnetic resonance imaging to quantify *in vivo* structural integrity of the locus coeruleus may provide biomarkers for noradrenergic experimental medicines studies in PSP.

## Introduction

The locus coeruleus is the principal source of noradrenaline, with diverse influences on arousal, behaviour, movement, and cognition [4]. The locus coeruleus is vulnerable to neurodegeneration in several diseases including Alzheimer’s disease, Parkinson’s disease, and progressive supranuclear palsy (PSP) [1, 10, 25, 30, 47, 52]. Here, we focus on the neuropathology of the locus coeruleus in PSP, a complex parkinsonian syndrome characterised by postural instability, falls, oculomotor impairment, cognitive, and behavioural changes. We test the hypotheses that in patients with clinical and pathologically confirmed diagnosis of PSP, there is severe degeneration and tau-pathology in the locus coeruleus, and that the degree of noradrenergic cell loss relates to clinical severity.

The locus coeruleus is located in the posterior margin of the rostral pons, near the lateral floor of the fourth ventricle. Despite its small size, with only tens of thousands of neurons in adult humans [1, 30, 34, 36, 39, 45], the locus coeruleus sends widespread projections to the neocortex, thalamus, and sub-cortical areas, sparing the majority of the striatum. Its connectivity enables a concerted release of noradrenaline in multiple target areas with modulatory effects on several physiological and cognitive functions including arousal, vigilance, sleep, attention, working-memory, and adaptive behaviour (reviewed by [4, 13]). Many of these functions are affected by PSP, over and above the PSP’s classical movement disorder. The motor and cognitive symptoms of PSP are not relieved by standard anti-parkinsonian medications including dopaminergic therapies. However, there is evidence, from studies in Parkinson’s disease, that deficits associated with locus coeruleus’ degeneration can be partially restored by noradrenergic therapies [6, 29], raising the possibility of noradrenergic treatments in PSP.

At the neuropathological level, the degeneration of the locus coeruleus in PSP includes neuronal loss and the presence of neuronal and glial inclusions of hyperphosphorylated 4-repeat isoforms of microtubule-associated protein tau [43, 51]. In humans, the noradrenergic neurons of the locus coeruleus contain a pigment called neuromelanin, which is a by-product of noradrenaline synthesis. The de-pigmentation of the locus coeruleus in PSP is so extensive that it is visible on gross *post mortem* examination of the brain. Recently, the development of high-resolution magnetic resonance imaging (MRI) sequences, sensitive to the paramagnetic features of neuromelanin [42], has renewed the interest in developing biomarkers for assessing the *in vivo* degeneration of the locus coeruleus in neurodegenerative diseases including PSP [5]. However, before these MRI methods can be further developed, it is necessary to quantify the neuronal loss in the LC *ex vivo* and determine whether this pathology relates to other neuropathological aspects in PSP such as the proportion of tau-positive inclusions, and to clinical severity.

Therefore, we quantified the locus coeruleus neuropathology in complementary ways. First, we estimated the total number of pigmented neurons *post mortem* in PSP patients in relation to a group of controls of similar age. Second, we estimated the number of pigmented neurons in the locus coeruleus that manifested neuronal inclusions comprising aggregated hyperphosphorylated tau. Third, we tested the correlations between pathological and clinical ratings. We confirm the severe loss of locus coeruleus neuron number, and a high rate of tau inclusions [20, 38], with a correlation between *ante mortem* disease severity (adjusting for time between latest clinical assessment and death), and the severity of neuronal loss in the locus coeruleus.

## Materials and methods

Brainstem tissue from patients and controls was obtained through the Cambridge Brain Bank at the Cambridge University Hospitals NHS Trust, UK (under the ethically approved protocol for “Neurodegeneration Research in Dementia”) and normative cognitive data from the PiPPIN cohort (“Pick’s disease and progressive supranuclear palsy prevalence and incidence study” [16]). Thirty-one patient donations were received between 2010 and 2017 from patients with a clinical and pathological diagnosis of PSP. The available fixed tissue blocks for two PSP-cases did not include the entire locus coeruleus so for these two we only report their percentage of pigmented neurons positive for tau-inclusions. Moreover, in two other cases, no immunohistochemistry could be performed because tissue sections detached from the glass slide.

The neuropathological diagnosis of PSP was based on the National Institute of Neurological Disorders and Stroke (NINDS) criteria. Clinical diagnoses were made according to the revised MDS 2017 criteria (Höglinger *et al*., 2017), based on the final clinical review (see Gazzina et al. for details [23]). This led to diagnoses of probable PSP-Richardson’s syndrome in n=22, possible PSP with predominant speech/language disorder (PSP-SL) in n=2, and possible PSP with predominant corticobasal syndrome (CBS) in n=7.

Many of the PSP patients had also participated in longitudinal cohorts studies at the Cambridge Centre for Frontotemporal Dementia and Related Disorders [16, 32]. Clinical, cognitive, and behavioural assessments included the PSP rating scale (PSPRS) [24], revised Addenbrooke’s cognitive examination (ACE-R) [35], Mini Mental State Examination (MMSE) [21], and revised Cambridge Behavioural Inventory (CBI-R) [48]. The PSPRS was available within 3 years of death for n=19 cases of PSP-Richardson’s syndrome, n=2 PSP-CBS and n=1 PSP-SL. For control tissue, we examined six Brain Bank donors with no history of neurological or psychiatric illness and of similar age and sex as PSP cases. *Ante mortem* clinical and cognitive data for control brain donors were not available. However, we compared the cognitive profile of the PSP patients to a control population of elderly individuals (n=60) recruited in a regional epidemiological study [16].

Group ages, brain weight, and cognitive scores were compared with two-samples independent t-test (Table 1 and 2). Sex differences between groups were tested using Fisher’s exact test (Table 1).

**Table 1.** Demographic and clinical characteristic of patients with PSP, and pathology-controls.

|  | **PSP**<br>**(n=31)** | **Controls**<br>**(n=6)** | **p-value** |
| --- | --- | --- | --- |
| Age (years) at Death |  |  |  |
| Mean ± SD | 75.4 ± 7.4 | 70 ± 5.6 | n.s.<sup>a</sup> |
| (Range) | (63 – 90) | (62 – 77) |  |
| Number (%) of males | 19 (61%) | 2 (33%) | n.s.<sup>b</sup> |
| Age (years) at symptom onset |  |  |  |
| Mean ± SD | 71.5 ± 8.3 | --- | --- |
| (Range) | (51 – 86) |  |  |
| Disease duration (years) |  |  |  |
| Mean ± SD | 6.9 ± 2.9 | --- | --- |
| Range | (3.1 - 15.5) |  |  |
| Brain weight (g) |  |  |  |
| Mean ± SD | 1196 ± 142.7 | 1222 ± 47.8 | n.s.<sup>a</sup> |
| Range | (960 - 1500) | (1153 – 1263) |  |

PSP, progressive supranuclear palsy. SD, standard deviation. n.s. not significant, $p > 0.05$ uncorrected,
by <sup>a</sup> Student's t-test or <sup>b</sup> Fischer's exact test

**Table 2.** Demographic and cognitive characteristics of the PSP patients and healthy controls from the “PiPPIN” regional epidemiological study.

|  | **PSP patients**<br>**(n=31)** | **Controls from**<br>**PiPPIN study**<br>**(n=60)** | **p-value** |
| --- | --- | --- | --- |
| Age at testing | 73.3 ± 7.7 | 71.4 ± 6.0 | n.s.<sup>a</sup> |
| Number (%) of males | 19 (62%) | 30 (50%) | n.s.<sup>b</sup> |
| PSPRS (PSP n=23) |  |  |  |
| Mean ± SD | 48.9 ± 14.1 | 0.32 ± 0.99 | t<sub>45</sub> = 17.2, p = 2.2e<sup>-21</sup> |
| Interval to death (year) |  |  |  |
| Mean ± SD | 1.8 ± 0.9 | --- | --- |
| ACER |  |  |  |
| Mean ± SD | 64.2 ± 16.0 | 95.7 ± 4.4 | t<sub>89</sub> = 14.3, p = 9.5e<sup>-25</sup> |
| CBI-R (PSP n=24) |  |  |  |
| Mean ± SD | 84.5 ± 52.7 | 6.8 ± 7.3 | t<sub>82</sub> = 11.3, p = 2.6e<sup>-18</sup> |
| MMSE |  |  |  |
| Mean ± SD | 21.9 ± 4.8 | 29.4 ± 0.9 | t<sub>89</sub> = -11.6, p = 1.4e<sup>-19</sup> |

PSP, progressive supranuclear palsy. PSPRS, PSP rating scale. ACER, revised Addenbrooke’s cognitive examination. CBI-R, revised Cambridge Behavioural Inventory. MMSE, Mini Mental State Examination, SD- standard deviation. n.s. not significant, p>0.05, by <sup>a</sup> Student’s t-test or <sup>b</sup> Fischer’s exact test. Subscript to t is degrees of freedom. Uncorrected p-values.

### Tissue processing and immunohistochemistry

The brain was removed at autopsy and the left cerebral hemisphere, left hemi-brainstem, and left cerebellum were fixed in 10% neutral buffered formalin for 2-3 weeks. Immunohistochemistry was performed on serial sections from brainstem, subcortical, and cerebral regions to identify the characteristic protein aggregates and pathological features of PSP and confirm the diagnosis *post mortem*. The initial diagnostic immunohistochemistry had used antibodies targeting Tau11/57 (from 2010 to mid-2016), or AT8 (from mid-2016 to 2017; MN1020, Thermo Scientific, USA). To exclude significant co-pathologies, additional immunohistochemistry included beta-amyloid (Clone 6F/3D, M0872, Dako, Denmark), alpha-synuclein (SA3400, Enzo life sciences, USA) and TDP-43 (TIP-PTD-P02, Cosmo Bio Co LTD, Japan).

To ensure that the entire locus coeruleus was available for the estimation of numbers of neurons, the diagnostic blocks containing the pons and lower midbrain were retrieved and added to the fixed samples containing the locus coeruleus. Brain tissue were embedded in paraffin and cut at 10 μm on a rotary microtome and mounted on SuperFrost Plus^™^ Adhesion Microscope Slides (J1800AMNZ, ThermoFischer Scientific, USA). Sections containing the locus coeruleus were immunostained for hyperphosphorylated tau (AT8) to visualise pathological tau aggregates. A series of 1 per 100 sections covering the entire locus coeruleus were deparaffinised in xylene and hydrated in progressively decreasing concentrations of alcohol and rinsed in running water. Antigen retrieval was performed at room temperature by incubating for 5 min in formic acid (33015-2.5L-M, Sigma-Aldrich, Germany), and blocked in 5% normal rabbit serum, before incubating with the primary antibody (AT8 1:500, MN1020, Thermo Scientific, USA) for 1h at room temperature. Sections were subsequently incubated with Polyclonal Rabbit Anti-Mouse Immunoglobulins/Biotinylated (E0354, Dako, Denmark) in 10% human serum and developed using the VECTASTAIN ELITE ABC Kit Avidin Biotinylated HRP Complex (PK-6100, VECTOR laboratories, USA) to yield a purple reaction product. Sections were counter stained with Vector Methyl green (H-3402-500, Vector laboratories, USA) to allow good visual discrimination between cell nuclei, tau aggregates, and endogenous neuromelanin (Figure 1).

**Fig. 1.**
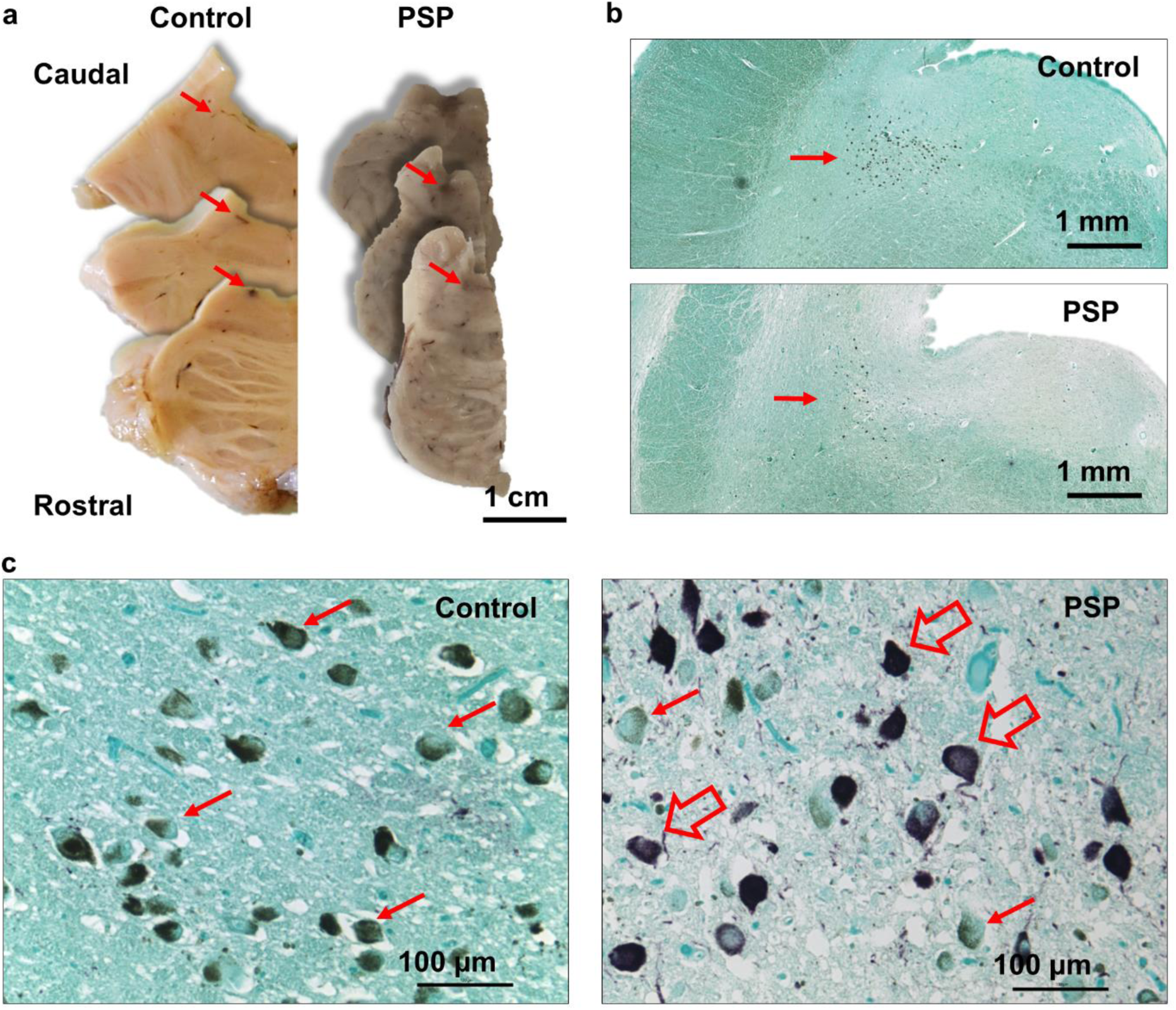
Representative images of locus coeruleus in control and PSP cases. (A) Photographs of pons sections including the locus coeruleus (arrows). (B) Micrographs showing the location of locus coeruleus near the floor of the 4^th^ ventricle in a control and PSP case (red arrows). Stitched images from micrographs taken using a 4X objective. (C) Micrographs taken using a 20X objective, showing pigmented neurons of the locus coeruleus (red arrows) and pigmented neurons with tau-inclusions (hollow red arrows). (B and C) Sections were stained for hyperphosphorylated tau (AT8) in purple using methyl green as counterstain, endogenous neuromelanin shown in brown.

### Quantification of pigmented neurons in locus coeruleus

A 2D design was used to estimate the total number of pigmented neurons (StereoInvestigator 11.0 64bit, Microbrightfield, USA) using an Olympus BX-53 microscope coupled with a Prior H128 computer-controlled x-z-y motorized stage and a high sensitivity Hitachi 3CCD video camera system. The locus coeruleus was outlined at 4x magnification and counting within 100% of that area was performed under a 20X objective. Pigmented neurons were defined by the presence of neuromelanin, which appears as brown granular staining within the cytoplasm of medium and large neurons in the locus coeruleus. When an AT8-positive aggregate was present in these pigmented neurons, the neuron was counted as tau immunoreactive. A neuron was counted when either the nucleolus or a well-defined nucleus was in focus (Figure 1).

The number of pigmented neurons was calculated by multiplying the number of neurons counted in the serial sections by 100 (the sampling frequency). As sections were 10 µm and we counted pigmented neurons in the entire cross-sectional area of locus coeruleus on each section, the height and area fractions were 1. The proportion of neurons with hyperphosphorylated tau-inclusions was determined by dividing the number of pigmented neurons positive for hyperphosphorylated tau-inclusions by the total number of pigmented neurons.

We used weighted least-square regression to examine the correlation between the number of pigmented neurons and PSPRS, ACER, MMSE, or CBI scores, and the relationship between the percentage of pigmented neurons with tau-inclusion and PSPRS score. To adjust for the difference in time between the last clinical assessment and death in relation to total disease duration, we included the time between clinical assessment and death as percent of disease duration (100*(1-”interval between testing and death”/”disease duration”)) (these values represented weights in the linear regression analyses). To test for an association between the number of pigmented neurons and the macroscopic scoring of locus coeruleus pallor, we used analysis of variance (ANOVA). Finally, Pearson’s correlations were used to test for associations between (i) the total number of pigmented neurons and the percentage of pigmented neurons with tau-inclusions and (ii) the pathological measures, disease duration, disease severity or other cognitive and neuropsychiatric measures. A p-value<0.05 was considered significant.

## Results

Table 1 summarises the clinical, demographic, and gross pathological characteristics of the brain donors. There were no significant differences in age, sex or brain weight between the PSP and pathology control cases. The control group from the PiPPIN epidemiological study was similar to the PSP group in terms of age and sex. However, as expected, PSP patients scored higher on the PSPRS and CBI, and lower on the ACE-R and MMSE than controls from the PiPPIN study (table 2).

The mean total number of pigmented neurons in the locus coeruleus was 50.6 x 10^3^ in controls (SD 1.4 x 10^3^) and 26.2 x 10^3^ in PSP patients (SD 1.4 x 10^3^). This 49% reduction of pigmented neurons in PSP patients was significant (t = 3.96, df = 33, p = 0.0004, Figure 2A).

**Fig. 2.**
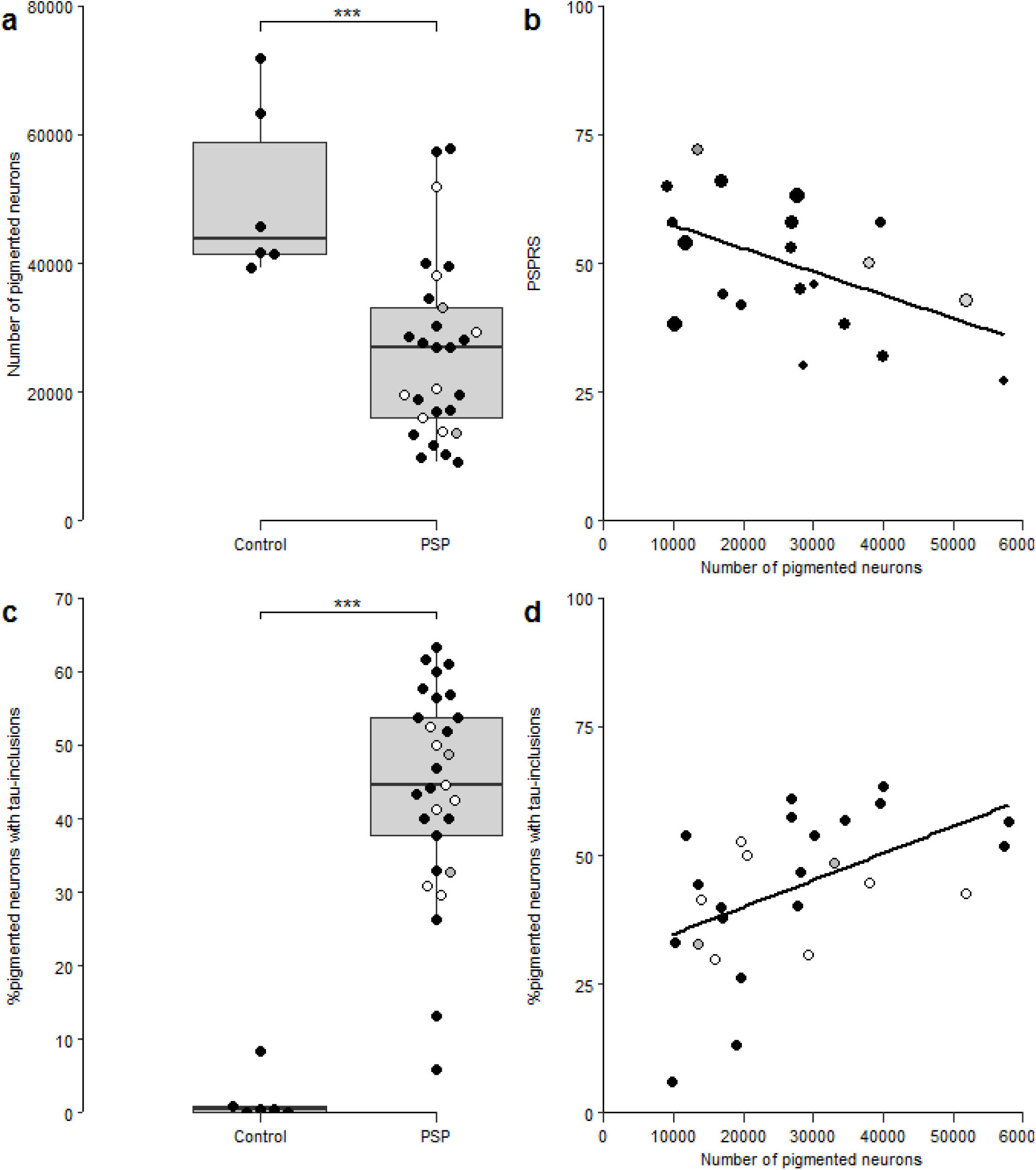
Loss of pigmented neurons in the locus coeruleus in PSP. Combined scatter and boxplots showing the number of pigmented neurons in the locus coeruleus in (A) controls and patients. (B) There was a negative correlation between the number of pigmented neurons and the PSP rating scale (PSPRS). The dot size for each individual scales with interval from assessment to death as a function of disease duration. (C) The percentage of pigmented neurons with tau-inclusions in controls and PSP. (D) The positive correlation between the number of pigmented neurons and number of pigmented neurons with tau-inclusions. Each dot represents an individual case, and for PSP cases (A-D), different colours of the dot represent specific PSP clinical phenotypes, i.e., black – probable PSP Richardson’s Syndrome, grey – possible PSP-SL, white – possible PSP-CBS. The lower and upper hinges of the grey boxes show the 25^th^ and 75^th^ percentiles, while the horizontal bar represent the median values. Whisker-plots display the range of data within 1.5 of the inter-quartile range, indicating that no extreme outlying values were observed. The linear regression lines are plotted in panel B and D.

Disease severity, as measured by total-PSPRS, was negatively correlated to the number of pigmented neurons (F(1,19)=5.9, t= −2.4, p = 0.026; W^-1/2^ * PSPSR = W^-1/2^ * 61.9 + W^-1/2^ *-4.5E-4*Neurons, where W is the weight matrix)(Figure 2B). The total number of pigmented neurons in the locus coeruleus did not correlate with disease duration (Pearson’s correlation, r = −0.22, p = 0.26) or age (Pearson’s correlation, r=-0.24, p = 0.16). We did not observe a significant correlation between the number of pigmented neurons and ACER (F(1,28) = 0.72, p = 0.40), MMSE (F(1,28) = 0.99, p = 0.33) or CBI (F(1,21) = 1.45, p = 0.24). The macroscopic scoring of locus coeruleus pallor was not significantly related to the microscopic counting of the number of pigmented cells (F(2,21)=1.37, p = 0.28).

The pigmented neurons positive for hyperphosphorylated tau were observed in four out of six controls, but the percentage of such neurons relative to total number of pigmented neurons was only 3% (range 0.5% to 9%). In PSP, the average percentage of pigmented neurons positive for hyperphosphorylated tau was 44% (range 6% to 63%). The increase in the number of pigmented neurons with tau-inclusions in PSP was significant (t=-7.26, df=33, p=2.5e^-8^)(Figure 2C). The percentage of pigmented neurons with tau-inclusions was not associated with PSPRS (F(1,19)=0.15, t = 0.39, p = 0.71). However, the percentage of neurons positive for hyperphosphorylated tau positively correlated with the total number of neurons in PSP patients (Pearson’s r = 0.51, p= 0.007)(Figure 2D).

## Discussion

This study demonstrates severe neurodegeneration in the locus coeruleus in PSP, with an average 49% reduction in the total number of pigmented neurons that are the principal source of noradrenaline to the brain. The total number of noradrenergic and neuromelanin-containing cells in PSP correlated negatively with disease severity, with fewer cells in those patients who had more severe disease in the last *ante mortem* clinical assessment. However, neuronal loss was not associated with age or disease duration, which suggests a possible role of the locus coeruleus degeneration in mediating clinical severity rather than simply reflecting an age-related effect or a consequence of a more prolonged disease course. The average percentage of pigmented neurons with hyperphosphorylated tau-inclusions in PSP was 44%, but this fraction was lower in PSP patients with more severe total neuronal loss. This suggests either a non-linear dynamic relationship between tau-aggregation and cell death, or the presence of a subset of pigmented neurons with low susceptibility to tau aggregation and death.

PSP pathology includes both neuronal and glia tau-inclusions, which are evident in the midbrain, pons, medulla, subthalamic nucleus, and globus pallidus, with additional susceptibility in some parts of the cerebral cortex such as the prefrontal and temporo-parietal gyri [17, 31, 51]. The pathological hallmarks of the locus coeruleus in PSP is depigmentation, neuronal tau-inclusions, and cell loss with gliosis. The locus coeruleus mediates a wide range of physiological, cognitive, and behavioural functions, via its noradrenergic innervation to forebrain structures; hence, the cell loss and tau pathology in the locus coeruleus is expected to have widespread clinical consequences in PSP [4, 49]. The average 49% loss of pigmented neurons that we found in this study is highly consistent with the 53% and 51% reduction of medium-to-large neurons in the locus coeruleus reported in two previous studies [38][33]. However, another semi-quantitative study, in which neurons positive for tyrosine hydroxylase were counted in single sections in the upper pons, did not report a significant loss of neurons in the locus coeruleus in PSP [20]. The difference across the studies may be due to the focus on the rostral pons rather than on the entire locus coeruleus, and to other possible differences in how the noradrenergic cells were defined and counted. In the present study, the neuromelanin-containing neurons were assumed to represent the entire noradrenergic neuronal population in the locus coeruleus. This is because the presence of the neuromelanin in the locus coeruleus cells is a consequence of excess catecholamines in the cytosol, which is in turn linked to the synthesis of the noradrenaline and its packing in synaptic vesicles [3, 4].

Another way to identify the LC cells producing noradrenaline is via the immuno-staining for tyrosine hydroxylase, the enzyme that converts L-tyrosine to L-DOPA before the conversion of L-DOPA to noradrenaline. Immunohistochemistry against the dopamine beta-hydroxylase, the enzyme that transform dopamine into noradrenaline, can also be used. These two biochemical markers are expressed by the same neuronal populations [14]: the noradrenergic cells that constitutes more than the 95% of neurons in the locus coeruleus [2, 38]. However, a proportion of tyrosine hydroxylase positive neurons, which are more numerous in the rostral than caudal pons, lack pigmentation [2, 14, 27]. The number of such neurons is limited in relation to the total neuronal number in the locus coeruleus [2, 14, 27]. Therefore, despite potential differences in the methods for identifying noradrenergic neurons, the findings from different pathological cohorts were in keeping with a significant reduction of noradrenergic neurons in PSP.

We observed considerable individual variability in the number of pigmented neurons in PSP, ranging from a normal level to one sixth of the control average. This variability in the number of pigmented neurons in PSP is in line with the variability seen macroscopically, although we did not find a direct relationship between the micro- and macro-scopic features. Currently, non-invasive methods to quantify the *in vivo* LC structural integrity, via the susceptibility of the MRI signal to the paramagnetic effects of neuromelanin, are emerging as promising tools for neurodegenerative disorders, especially at ultra-high field [42]. The association between *post mortem* depigmentation and the neuromelanin-related MRI signal in the LC may therefore provide support for *in vivo* biomarkers for noradrenergic dysfunction in PSP. This could facilitate the monitoring of the disease progression and treatment response [5, 40].

Our second hypothesis was that the loss of LC pigmented neurons relates to disease severity, measured via the PSPRS. This was confirmed. The PSPRS is the most widely used clinical scale of disease severity and progression in PSP in observational studies and clinical trials. It encompasses diverse domains of symptoms and signs that are typically endorsed by PSP patients and their carers, including behavioural, gait, mobility, bulbar, limb, and oculomotor problems. Other disorders and even normal aging can influence the total PSPRS score [24], but in the context of PSP clinical syndromes, the PSPRS has high internal consistency and replicability across different countries (e.g., there is a typical 10-12 points decline per year in Richardson’s syndrome) [7, 24]. The total score and domains of the PSPRS have been associated with PSP-related regional atrophy in the brainstem [18, 28, 46, 50] but not yet related to specific brainstem nuclei as the locus coeruleus [37] or *in vivo* tau pathology as measured via positron-emission tomography [19, 50].

Not all pigmented neurons in the locus coeruleus are affected equally by PSP. On average, 44% of the pigmented neurons had tau-inclusions. This accords with a semi-quantitative study showing tau inclusions in 42% of the noradrenergic neurons [20], and the presence of tau inclusions in 77% of the noradrenergic neurons counted using stereology [38]. The current cohort was larger than those employed in previous studies and found high variability, 6-63%, of LC neurons having tau-inclusions. This implies that differences between our findings and previous results might depend on the use of different sample sizes with variable estimates. Differences in the analytical procedures used or in the demographic characteristics of the cohorts may also have contributed to the different percentages of tau-positive neurons across studies. We found a significant positive correlation between the percentage of pigmented neurons with tau-inclusions and the total number of pigmented neurons. However, the percentage of the pigmented neurons was not associated to tau pathology or disease severity. These results are consistent with the hypothesis that the accumulation of tau precedes and contributes to the neuronal death [49], with differential susceptibility to cell death with tau-positive neurons as the disease progresses, or as a result of subtypes of pigmented neurons.

The association between the locus coeruleus integrity and disease severity raises the possibility of noradrenergic treatment strategies. One study reported that alpha 2 noradrenergic receptor antagonism did not alleviate motor symptoms in PSP [41]. However, there is more favourable evidence, from studies in Parkinson’s disease (PD), that the noradrenalin reuptake inhibitor atomoxetine can improve motor control and executive functions in a subset of patients [6, 29, 53]. Clinical trials of noradrenergic treatments are warranted in PSP [40]. However, the relationship between the loss of pigmented neurons in the locus coeruleus and disease severity calls for a stratified approach in clinical trials, which might exploit emerging imaging biomarkers of LC structural integrity.

The current data can also be compared to those reported in the tauopathy of Alzheimer’s disease, in which the neurofibrillary tangles in the locus coeruleus are present at the pre-symptomatic stages I and II [8, 9, 11, 30, 45], although the neuronal loss in the locus coeruleus becomes more severe from Braak stage III onwards [1, 11, 30, 45]. The cell loss in the locus coeruleus is associated with global clinical severity in Alzheimer’s disease [1, 30]. In contrast to the correlation between the percentage of pigmented neurons with tau-inclusions and neuronal loss we observed in PSP, the percentage of noradrenergic neurons with intracellular neurofibrillary tangles does not correlate with the loss of locus coeruleus neurons in Alzheimer’s disease [11]. This difference may depend on the different tau isoforms present in AD and PSP. Pre-clinical studies have indicated that the relationship between tau-aggregation and neuronal loss in the locus coeruleus may be not linear, even though the overexpression of tau is sufficient to induce tau-inclusions and neuronal death in the locus coeruleus [12]. From human studies, there is conflicting evidence on whether tau deposition impairs the expression of tyrosine hydroxylase and the capacity for synthesizing noradrenaline in Alzheimer’s disease before neuronal death in the locus coeruleus [22, 45]. The same seems to true in PSP, as shown by a recent study which reported more severe loss of neurons staining positive for tyrosine hydroxylase than the total number of neurons [38]. However, abnormal tau deposition may be exacerbated by the loss of the noradrenergic neurons in animal models of Alzheimer’s disease [26] and genetic frontotemporal dementia [12]. Future studies addressing the causal relationships between tau inclusions, neuronal loss, noradrenergic signalling in the locus coeruleus in PSP and other neurodegenerative disorders will help to refine future interventional studies.

The present study has several limitations of this study. First, as the selection of cases was based on the *post mortem* neuropathological diagnosis of PSP, we lack the PSP rating scale in some cases with clinical diagnosis of PSP-CBS. Second, as the PSPRS was necessarily obtained *ante mortem*, the interval between clinical and cognitive testing and death varied between patients, although we sought to mitigate this issue by including, in our statistical models, the interval between the last assessment and death (in terms of percentage of total disease duration). The association between pigmented neurons and PSP rating scale remained significant with and without this weighting in the statistical model. Third, as we only focused on the locus coeruleus pathology in PSP, we cannot exclude *a priori* that the association between the PSP rating scale and locus coeruleus pathology reflect a more general association with neurodegeneration at secondary sites, including the targets of LC innervation. Fourth, in the present study we counted the pigmented cells in the locus coeruleus in serial sections selected using systematic sampling. We counted cells on the series of 2D sections rather than 3D which is necessary for a true unbiased stereological method. Our estimates are therefore not unbiased and may be skewed towards overestimation as the section thickness is less than the average diameter of pigmented cells [15]. Further, in a 2D fractionator larger particles will have a greater chance of being sampled [44], which may contribute further to an overestimation bias. We estimated the total number of pigmented neurons to 56,000 in the left locus coeruleus in controls. This estimate is larger than what has been reported in a number of stereological studies where estimate the unilateral number of pigmented neurons to range between 16,000-19,000 [1, 30, 34, 36, 39]. However, Theofilas et al. [45] used stereology to estimate the total number of mid-to large neurons in the locus coeruleus in 68 aged individuals from Braak-stage 0 –VI. They observed a mean of 46,500 mid-to large sized neurons in Braak stage 0, and a total range across all braak stages from 6,674 to 137,910 neurons. Differences in the stereological methodologies e.g. the physical vs. the optical disector or optical fractionator, which cell populations was counted e.g. pigmented [1, 34, 36, 39], tyrosine hydroxylase expressing [30] or size [45], and whether one counts the soma profile or nucleolus may contribute to the discrepancy between studies. Recognizing the inherent potential for biased estimates using a 2D fractionator design, we assume that this bias is constant across subjects and thus, does not invalidate the correlations between neuronal numbers and tau-load or disease severity.

To conclude, we provide evidence for severe degeneration in the locus coeruleus in PSP, and an association between the number of noradrenergic neurons containing neuromelanin and disease severity. However, neuronal loss does not appear to be a simple function of age, disease duration or the presence of hyperphosphorylated tau-inclusions. The use of the neuromelanin pigment to identify *post mortem* the noradrenergic cells in the LC also supports the development of non-invasive *in vivo* neuroimaging markers that exploit the MRI signal linked to the presence of neuromelanin [5]. The degeneration of the locus coeruleus may be relevant to the development of better stratification procedures in clinical trials. Our study also highlights the importance of considering noradrenergic restoration as a potential treatment for PSP.

## Data Availability

The datasets used are available from the corresponding author on reasonable request.

## List of Abbreviations

ANOVA: analysis of variance
ACE-R: revised Addenbrooke’s cognitive examination
CBI-R: revised Cambridge Behavioural Inventory
CBS: corticobasal syndrome
MRI: magnetic resonance imaging
MMSE: Mini Mental State Examination
NINDS: National Institute of Neurological Disorders and Stroke
PSP: progressive supranuclear palsy
PSPRS: PSP rating scale
PSP-SL: possible PSP with predominant speech/language disorder

## Declarations

## Acknowledgements

We thank the donors and their families.

## Ethics approval and consent to participate

The study ethics was approved by the Health Research Authority, NHS, England (IRAS-202 802, “Neurodegeneration Research in Dementia”). The PiPPIN (Pick’s Disease and Progressive Supranuclear Palsy: Prevalence and Incidence) Study was approved by Cambridge’s research ethics committee. The study was conducted in accordance with the 1964 Helsinki declaration.

